# Modeling the Impact of Pediatric RSV Immunization in Massachusetts, 2024–2025

**DOI:** 10.64898/2026.06.05.26354236

**Authors:** Laura Jones, Rosa Ergas, Andrew Tibbs, Elizabeth T. Russo, Josh Norville, Boudu Bingay, Catherine M. Brown, Nicholas G. Reich, Remy Pasco

**Affiliations:** Massachusetts Department of Public Health, Boston, Massachusetts, US; University of Massachusetts, School of Public Health and Health Sciences, Amherst, Massachusetts, USA; University of Colorado Boulder, Boulder, Colorado, USA

**Keywords:** Epidemiology, Immunization, Vaccination, Respiratory Syncytial Viruses, Pediatrics, Statistical Model, Mathematical Model, Epidemiological Models, Compartmental Models

## Abstract

**Background:** Pediatric immunizations for Respiratory Syncytial Virus (RSV), including monoclonal antibodies for infants and vaccines for pregnant people, have become broadly available and can prevent severe RSV outcomes in infants. However, quantifying the impact of RSV immunization in prevention of severe pediatric illness at the population-level is limited by lack of RSV case surveillance data. The Massachusetts Department of Public Health (DPH) conducted a modeling analysis using routine public health surveillance data to estimate the state-level impact of new RSV immunization products on Emergency Department (ED) visits and hospitalizations in Massachusetts for highest risk pediatric groups.

**Methods:** A scenario projection tool, called R.Scenario.Vax, was utilized to simulate RSV-associated ED hospital encounters by age group in the context of newly available immunizations. ED visit and hospitalization data from the National Syndromic Surveillance Program (NSSP) during the time period 10/08/2017–10/19/2024 were analyzed, scaled to account for changes in RSV testing practices over time and missing encounter volume in historic data, and utilized to inform model fit of a “typical” RSV season. RSV immunization data from the Massachusetts Immunization Information System (MIIS) for the 2023–2024 and 2024–2025 RSV seasons informed high and moderate pediatric RSV immunization coverage scenarios and their impact was compared to a counterfactual reference scenario of no new immunizations. Median projections were quantitatively and qualitatively compared to observed 2024–2025 season data. Percent reduction in hospital encounters and encounters averted per 10,000 population were calculated for each scenario as compared to the reference.

**Results:** Projections for the youngest at-risk age groups showed significantly lower RSV-associated ED visits and hospitalizations during the 2024–2025 season for both high and moderate immunization coverage scenarios. Median projections for infants under 6 months old in the highest coverage scenario, wherein nearly all infants were immunized, showed 72.6% lower ED visits and 73.4% lower hospitalizations when compared to the reference scenario, equating to 262 ED visits and 85 hospitalizations averted per 10,000 population.

**Conclusions:** Our results support the use of modeling methods for public health insights and suggest that RSV immunizations for infant populations result in significantly lower RSV-related ED encounters in Massachusetts.

## Introduction

Respiratory Syncytial Virus (RSV) is a common respiratory virus that can lead to severe illness, including hospitalization and death, especially among young children and older adults. Data about RSV cases are not universally reportable to public health jurisdictions in the United States. As a result, insights about the impact and burden of RSV through traditional infectious disease surveillance activities conducted by state and local public health jurisdictions can be limited. In Massachusetts, emergency department (ED) visits and hospital admissions captured in the state’s National Syndromic Surveillance Program (NSSP) data [1] are a primary measure of disease burden associated with overall respiratory illness and provide near real-time awareness of hospital utilization, an important measure of disease burden associated with severe RSV infection. However, syndromic surveillance data are deidentified and not easily integrated with other data sources, such as person-level immunization records. This limits the ability to measure directly how RSV immunization prevents disease in real-world contexts.

Historically, RSV cycled in an annual, seasonal pattern, but trends in RSV patient volume and seasonal timing have shifted in recent years, due to disease transmission disruptions caused by the COVID-19 pandemic [2,3]. Recent literature indicates that RSV testing practices at hospitals changed over time, both pre- and post-pandemic, further complicating understanding of disease trends from traditional surveillance data [4,5]. Furthermore, opportunities for immunization have emerged in recent years, and have influenced trends in severe illness for RSV [6]. Beginning in 2023, new RSV immunization products became available for use by at-risk groups, including a vaccine for pregnant people and the monoclonal antibody *nirsevimab* approved for routine use for infants, both aimed at preventing severe disease in infants and young children (Supplement). RSV vaccines also became available in 2023 for adults aged 65 years and older. *Nirsevimab* for infants was limited in the 2023–2024 respiratory season due to a supply-demand incongruity but was more widely available in the 2024–2025 season. Indications are that there has been a gradual return to pre-pandemic seasonal disease patterns [2], but expansion beyond conventional public health surveillance methods for RSV can offer additional insights about disease trends and the impact of new public health interventions.

The use of advanced mathematical models for characterization and prediction of infectious disease trends–has expanded the possible depth of epidemiological analyses, as modeling can account for complex disease and behavior dynamics not directly observable or inferable through conventional surveillance data sources and statistical methods [7–9]. However, the use of modeling at the level of state, tribal, local, and territorial (STLT) public health agencies is not ubiquitous and is often hampered by resource constraints, lack of experience and expertise with modeling, and other operational and administrative barriers. More accessible “off-the-shelf” modeling tools are now available to STLT public health jurisdictions, designed to help inform rapid public health decision-making and applied epidemiologic approaches for prevailing public health concerns and emerging threats.

The Massachusetts Department of Public Health (DPH) utilized a scenario projection tool, called R.Scenario.Vax [10], to simulate RSV-associated hospitalizations in the context of newly available immunizations to support public health surveillance, communication, and decision-making, and to demonstrate the utility of modeling tools for applied public health. To assess the feasibility and practicality of utilizing a scenario projection modeling, the tool was piloted during the 2024–2025 RSV season using historical syndromic surveillance and immunization data inputs and preliminary estimates of 2024–2025 season data as a reference for model inputs. Work focused on infant populations given their burden of severe illness. The role of changing RSV testing practices on surveillance data was also considered for broad-stroke adjustments to the surveillance data to enhance public health insights.

## Methods

### RSV ED Visit and Hospitalization Data

NSSP ED visit and hospitalization data (encounters that resulted in hospital admission) provided measures of severe outcomes for RSV. RSV-associated ED encounters and hospitalizations, defined as patient encounters categorized with the “CDC Respiratory Syncytial Virus v1” syndrome definition [11] at Massachusetts hospitals for patients with Massachusetts-based addresses, were included in this analysis. Historical RSV season data inputs used to inform model fit between 10/08/2017–10/19/2024 were analyzed. Descriptive summaries were generated by age group (<6 months, 6–11 months, 1–4 years, 5–64 years, 65–74 years, and ≥75 years) and season (epidemiologic weeks 41 through 22) for ED visits and hospitalizations. Encounters missing age data were excluded from analysis (2.1% of ED visits (n=1,067); 0.3 % of hospitalizations (n=45)).

To calculate a “typical” RSV season using the modeling tool, historical data prior to January 1, 2019 were adjusted to account for missing encounter volume from one large hospital system not yet enrolled in NSSP; ED visit and hospitalization data were scaled using the 2019–2020 RSV season percentage of statewide visits for that hospital system by age group (Supplement). To account for the impact of changes in RSV testing practices over time, historical data were then scaled by the change in encounter volume between 2018–2019 and 2023–2024 RSV seasons by age group (Supplement) [4,12]. Lastly, the 2017–2018 and 2018–2019 season historical data were also scaled to account for year-over-year increase in respiratory testing practices observed prior to the COVID-19 pandemic (Supplement) [5]. These scaled data were then aggregated into a weekly time series for input into the modeling tool; weeks after April 1, 2020, were excluded from the model due to the disruptions in disease trends during the COVID-19 pandemic. Descriptive summaries by age group were also generated for the observed 2024–2025 RSV season (October 12, 2024–May 31, 2025) for both ED visits and hospitalizations as reference for model output and evaluation.

To represent a “typical” season’s encounter burden by age group, overall age distribution for both ED visits and hospitalizations, were calculated using the average proportion of encounters by age group from the prior two RSV seasons (2022–2023 and 2023–2024) to account for the most recent trends in age of patients with ED visits or hospitalizations for RSV (Supplement).

### Immunization Data and Modeling Scenarios

Preliminary estimates of 2024–2025 RSV immunization coverage from the Massachusetts Immunization Information System (MIIS) as of April 30, 2025, were utilized to inform realistic scenarios of interest to support the pilot of this tool during the 2024–2025 season. Infant immunizations administered “at birth” were defined as distinct patients given monoclonal antibodies under 1 month of age during the 2024–2025 season. “Catch-up” doses of monoclonal antibodies were those administered to infants born prior to October 1, 2024, who were still less than 8 months old during the 2024– 2025 season. Immunizations given during pregnancy included RSV vaccinations administered to any individual in MIIS ages 5 through 49, regardless of reported sex. To estimate overall impact of RSV immunization on adult hospitalizations during the 2024– 2025 season, historical RSV vaccination data for older adult age groups (65–74 years, 75+ years) for the 2023–2024 season from MIIS were also included in the model scenarios.

Using the preliminary 2024–2025 immunization coverage numbers and an estimated 70,000 live births each year in Massachusetts [13], we constructed possible scenarios to explore the impact of A) high RSV immunization coverage with only monoclonal antibody administration to infants, B) high coverage with use of both monoclonal antibodies and vaccinations administered during pregnancy, C) moderate coverage most similar to the observed values during the actual 2024–2025 season, and lastly, a reference scenario serving as the comparison that included no new immunizations in the 2024–2025 season (Table 1). High coverage scenarios were designed to explore the impact of immunizing nearly all infants born in Massachusetts each year, through either the recommended monoclonal antibody administration or vaccination during pregnancy. The high coverage scenarios also included a hypothetical older adult population cumulative coverage of approximately 73%, to reflect the equivalent of the highest seasonal influenza vaccination coverage observed in those age groups.

**Table 1.** RSV Immunization Coverage Scenarios of Interest, 2024–2025 RSV Season, Massachusetts, United States.

| Assumed Doses Administered (% coverage), by type and age group |  | Immunization Coverage Scenarios of Interest |  |  |  |
| --- | --- | --- | --- | --- | --- |
|  |  | Scenario A: High coverage, with infant monoclonal antibody focus | Scenario B: High coverage, with some adult vaccinations administered during pregnancy | Scenario C: Moderate coverage, most similar to actual 2024–2025 season coverage | Reference Scenario: No new immunizations in 2024–2025 |
| Monoclonal antibodies | At birth | 24,000 (34.3%) | 18,000 (25.7%) | 11,000 (15.7%) | . |
|  | Catch-up | 46,000 (65.7%) | 36,000 (51.4%) | 23,000 (32.9%) | . |
| Vaccination during pregnancy | All ages | . | 16,000 (22.9%) | 10,000 (14.3%) | . |
| Older adult vaccination* | 65–74 yrs | 387,000 (53.9%) | 387,000 (53.9%) | 36,000 (5.0%) | . |
|  | 75+ yrs | 250,000 (48.7%) | 250,000 (48.7%) | 50,000 (9.7%) | . |

Percent coverage of monoclonal antibody and vaccinations administered during pregnancy based on an estimated 70,000 live births each year in Massachusetts. Source: Massachusetts Department of Public Health, Registry of Vital Records and Statistics [13]. \*All scenarios included RSV vaccination coverage of older adult populations received prior to or during the 2023–2024 respiratory season (for ages 65–74 years old: 137,520 doses (19.1% coverage); for ages 75+: 125,371 doses (24.4% coverage)). Source: Massachusetts Immunization Information System [14]. Percent coverage population denominators based on 2020 UMass Donahue Institute population estimates [15].

### Modeling Analysis

All analyses were performed using R, version 4.4.1, and RStudio, version 2024.09.1+394 [16,17]. The R.Scenario.Vax package [10] was used to model the scenarios and generate projections. The modeling tool utilizes an age-structured compartmental transmission model with fixed susceptibility and infectiousness parameters based on published literature, and incorporates relevant census data to accommodate birth rates, net migration, and age-specific population distribution [10]. Model parameters for RSV infant immunization effectiveness against ED visits and hospitalization were based on data from clinical trials and observational studies available at the time of analysis; effectiveness was set at 80% for monoclonal antibody *nirsevimab* and 57% for immunity obtained through vaccination during pregnancy [18–23] (see Supplement for full list of model parameters). Using maximum likelihood estimation, the model was fitted to the scaled historical time series and age distribution data. Projections were generated for the specified immunization scenarios for the period October 1, 2024, through June 1, 2025, and simulated 100 trajectories. The median and 2.5% and 97.5% quantiles from the trajectories were provided as 95% projection intervals (PIs) [10,12].

Pearson correlation coefficients were calculated for the median projection estimates of Scenario C (closest immunization coverage to the actual season) with the observed 2024–2025 RSV season values to capture the agreement in trend. Moving epidemic method intensity thresholds calculated by DPH for the 2024–2025 RSV season were also qualitatively referenced to further contextualize Scenario C model outputs. To summarize model outputs, two measures were calculated based on the 100 simulated trajectories. First, percent reduction in hospital encounters between the reference scenario and each immunization scenario was calculated as [(total reference scenario encounters – total immunization scenario encounters) / total reference scenario encounters] x 100 for all ages and stratified by pediatric at-risk age group. Second, the estimated rate of infant ED visits and hospitalizations averted per 10,000 population was calculated by subtracting the projections for each scenario from the reference scenario projection estimates, with the population denominator an estimated 70,000 MA live births [13]. For both measures, we report the median across the 100 simulations, with 95% uncertainty intervals (UIs) calculated as the 2.5th and 97.5th percentiles of the simulated distributions to capture the range of plausible outcomes given model parameters and assumptions.

## Results

### Historical RSV Hospital Encounter Data

Between October 8, 2017, and October 19, 2024, 51,744 ED visits related to RSV were observed, including 14,785 hospitalizations (Table 2). The number of historical hospital encounters by RSV season ranged from 212 to 13,092 for ED visits and 35 to 3,516 for hospitalizations, with the highest volume of encounters seen in the two most recent historical seasons (2022–2023 and 2023–2024). Infant populations less than 1 year of age had the highest volume of ED visits (a combined total of 18,695 encounters). Infants less than 6 months old had the highest proportion of hospitalizations (29%), followed by the 1–4-year-olds (22%), and those over 75 years old (18%) (Table 2).

**Table 2.** Count of Historical RSV Emergency Care Encounters, 10/08/2017–10/19/2024, Massachusetts, United States.

|  | *Type of Hospital Encounter* |  |
| --- | --- | --- |
|  | *ED Visits* | *Hospitalizations* |
| Total encounters | 51,744 | 14,785 |
| Missing age | 1,067 | 45 |
| RSV Season* |  |  |
| 2017–2018 | 3,182 | 987 |
| 2018–2019 | 4,673 | 1,600 |
| 2019–2020 | 6,463 | 2,206 |
| 2020–2021† | 212 | 35 |
| 2021–2022‡ | 4,429 | 1,143 |
| 2022–2023‡ | 13,092 | 3,465 |
| 2023–2024‡ | 11,509 | 3,516 |
| Age Group (%) |  |  |
| <6 months | 11,241 (22%) | 4,201 (29%) |
| 6–11 months | 7,454 (15%) | 1,611 (11%) |
| 1–4 years | 18,010 (36%) | 3,210 (22%) |
| 5–64 years | 8,265 (16%) | 1,948 (13%) |
| 65–74 years | 2,066 (4%) | 1,160 (8%) |
| 75+ years | 3,641 (7%) | 2,610 (18%) |

Abbreviations: ED, Emergency Department. \*RSV Season = October through May (epidemiologic weeks 41 through 22). †The 2020–2021 season was associated with a significant decrease in patterns of disease transmission due to the COVID-19 pandemic. ‡Increase in post-pandemic RSV-associated hospital encounters has been partially attributed to changes in respiratory testing practices that impact hospital coding practices [4,5]. Source: Bureau of Infectious Disease and Laboratory Sciences, Syndromic Surveillance program.

Qualitative assessment of scaled historical seasons’ hospital encounters showed more consistent peak seasonal volume over time for ED visits than was observed in the unscaled data. Differences in peak volume remained for hospitalizations between the three historical seasons after scaling was applied (Figure 1).

**Figure 1.**
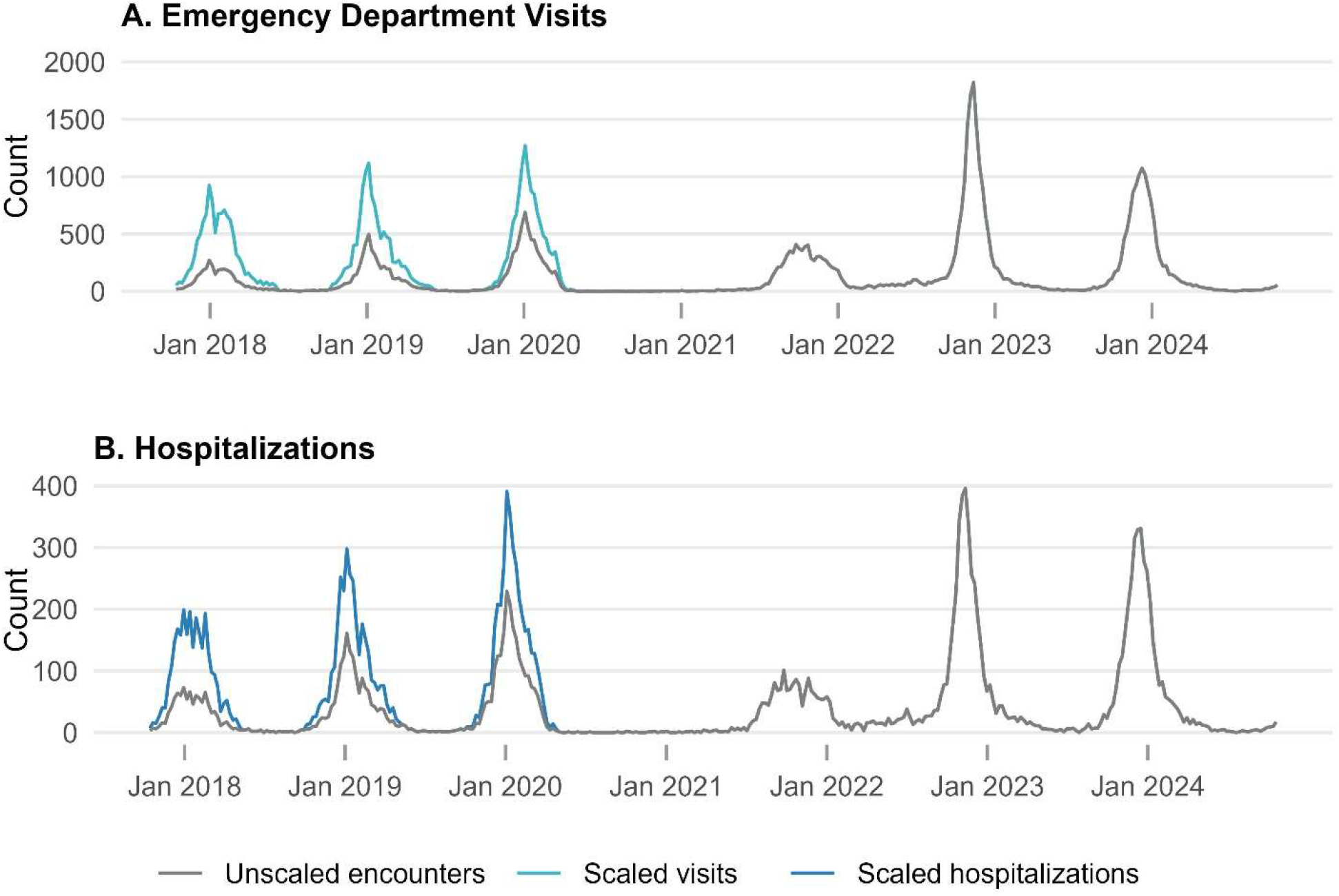
Scaled weekly RSV Emergency Department visits and hospitalizations, 10/08/2017– 10/14/2024, Massachusetts, United States. ED encounter scaling included adjustments for missing facility volume and to account for changes in RSV testing practices over time (both pre- and post-pandemic). Source: Bureau of Infectious Disease and Laboratory Sciences, Syndromic Surveillance program.

### Scenario Projections for the 2024–2025 Season

Scenario projections of total hospital encounters across all age groups showed the lowest volume of ED visits and hospitalizations in Scenario A (high immunization coverage, with a focus on monoclonal antibodies for infant age groups) followed by Scenario B (high immunization coverage, with both monoclonal antibody and vaccination during pregnancy focus) (Table 3). Observed ED visits and hospitalizations for the 2024–2025 season were higher than the median projections of all scenarios; qualitative assessment of the seasonal intensity thresholds calculated by DPH for the 2024–2025 RSV season indicated the peaks of both ED visits and hospitalizations for RSV were categorized as high when compared to previous seasons [24] (Supplement). Observed hospital encounters for all ages fell within the 95% projection intervals. Pearson correlation coefficients for Scenario C median projection estimates (closest immunization coverage to the actual season) showed strong positive correlations with the observed 2024–2025 RSV season values of r(32) = 0.940 (95% CI, 0.882-0.970), *P* < .001) for ED visits and r(32) = 0.962 (95% CI, 0.924-0.981], P < .001) for hospitalizations (Supplement).

**Table 3.** Modeled Scenario Projections and Observed Count of RSV Emergency Department Encounters, 2024–2025 Season, Massachusetts, United States.

|  | 2024–<br>2025<br>Season | 2024–2025 Scenario Projections* |  |  |  |
| --- | --- | --- | --- | --- | --- |
|  | Observed | Scenario A | Scenario B | Scenario C | Reference<br>Scenario |
| Emergency Department Visits, count (95% PI) |  |  |  |  |  |
| All ages | 10,778 | 7,782<br>(3,375–<br>13,846) | 8,007<br>(3,483–<br>14,200) | 9,019<br>(3,948–<br>15,858) | 10,267<br>(4,520–<br>17,850) |
| Hospitalizations, count (95% PI) |  |  |  |  |  |
| All ages | 3,027 | 2,085<br>(906–3,745) | 2,157<br>(940–3,857) | 2,593<br>(1,139–4,579) | 3,015<br>(1,332–5,263) |

Abbreviations: PI, projection interval. \*Scenarios described in Table 1.

Projected hospital encounters for at-risk pediatric age groups were similarly lowest for Scenario A followed closely by Scenario B (Figure 2). Projected hospital encounters for infants less than 6 months old in Scenario C, reflecting immunization coverage most similar to that observed for the 2024–2025 season, were considerably lower than the counterfactual scenario (Figure 2).

**Figure 2.**
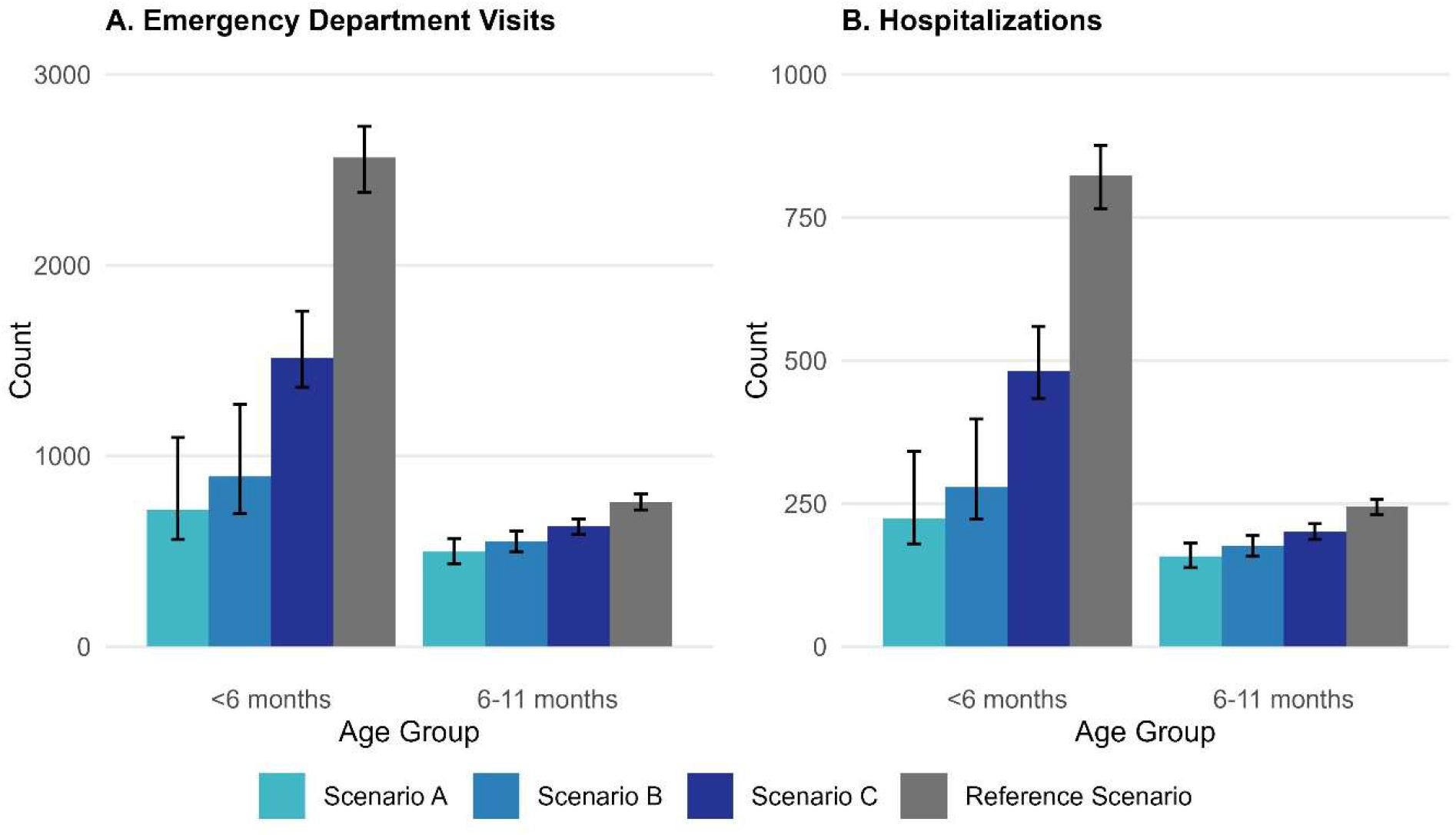
Projected RSV hospital encounters by at-risk pediatric age group, 2024–2025 season, Massachusetts, United States. 95% Projection Intervals included as error bars. Scenarios described in Table 1.

Compared to the median reference scenario (no new immunizations administered), ED visits were projected to be 72.6% lower (95% UI, 57.6%-77.6%) in the highest coverage scenario (Scenario A) and 65.8% lower (95% UI, 51.0%-72.1%) in the second highest scenario (Scenario B) for the youngest age group (Table 4). Infants less than 6 months old also showed 73.4% lower (95% UI, 59.0%-77.6%) and 66.7% lower (95% UI, 52.3%-72.3%) hospitalizations for Scenarios A and B, respectively. The findings for the high coverage scenarios for those under 6 months old correspond to between 238 (95% UI, 187-277) and 262 (95% UI, 211-301) ED visits averted per 10,000 population and between 77 (95% UI, 62-89) and 85 (95% UI, 70-97) hospitalizations averted per 10,000 population. In the scenario most similar to the observed 2024–2025 immunization coverage, infants in that age group saw 41.4% lower (95% UI, 32.1%-45.3%) ED visits and 41.9%-lower (95% UI, 32.9%-45.4%) hospitalizations compared to the median reference scenario projection, corresponding to 150 visits (95% UI, 118-174) and 49 hospitalizations (95% UI, 39-56) averted per 10,000 population. Lower ED visits and hospitalizations were also seen among the 6–11-month-old age group, demonstrating the impact of their partial eligibility for catch-up doses and/or partial conferred immunity during pregnancy; for the highest coverage scenarios, ED visits were 27.7% (95% UI, 20.2%-33.6%) to 34.7% (95% UI, 25.4%-41.9%) lower, and hospitalizations were 28.3% (95% UI, 20.8%-34.1%) to 35.5% (95% UI, 26.1%-42.4%) lower. The high coverage scenarios reduction in hospital encounters across all ages showed the population-level impact of immunizations in all at-risk populations: 23.9% lower (95% UI, 19.7%-26.4%) ED visits and 30.7% lower (95% UI, 25.8%-32.9%) hospitalizations than the reference.

**Table 4.**
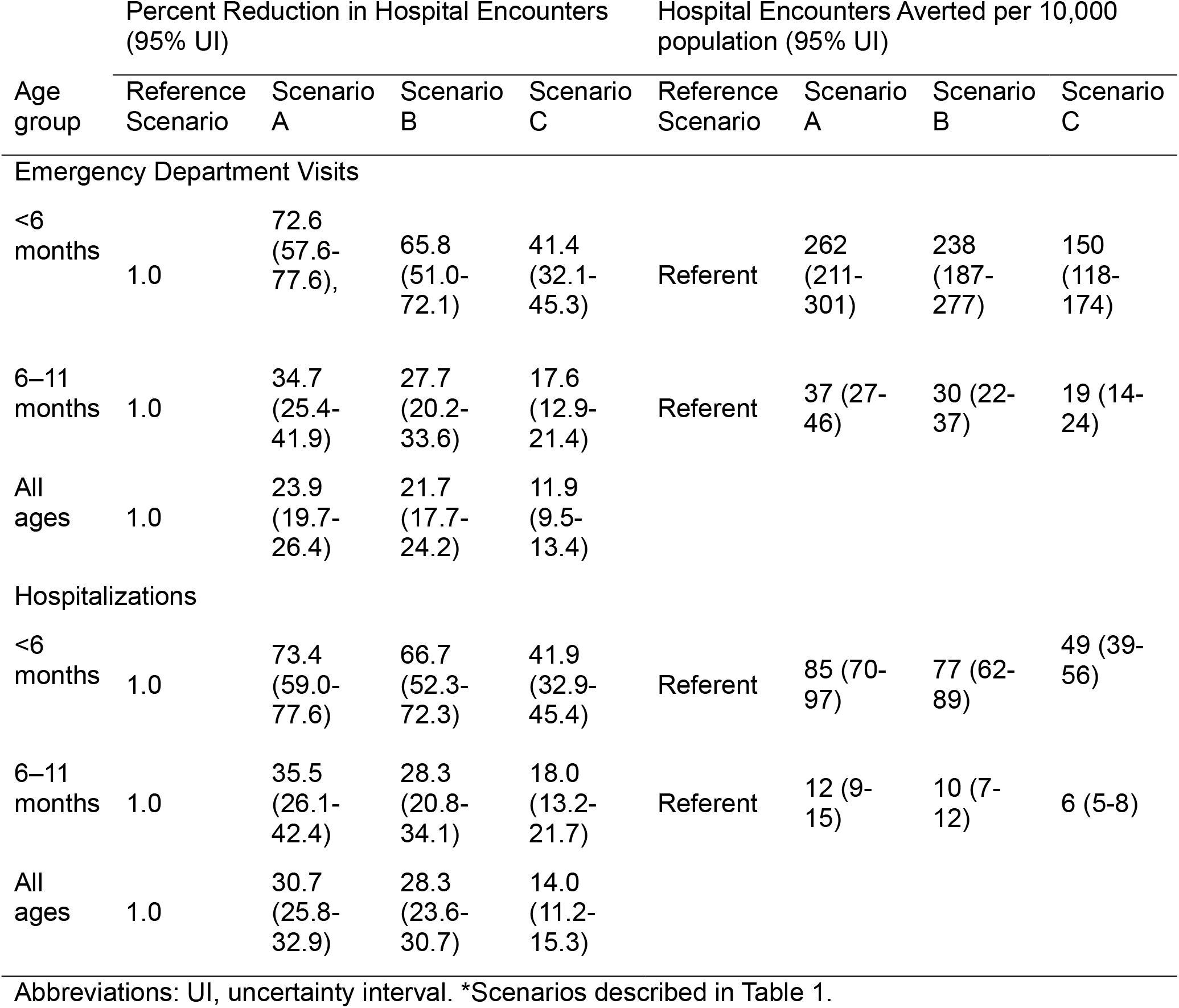
Projected Percent Reduction in RSV Hospital Encounters and Encounters Averted, Immunization Coverage Scenarios vs. Reference Scenario, 2024–2025 Season, Massachusetts, United States.

| Age group | Percent Reduction in Hospital Encounters (95% UI) |  |  |  | Hospital Encounters Averted per 10,000 population (95% UI) |  |  |  |
| --- | --- | --- | --- | --- | --- | --- | --- | --- |
|  | Reference Scenario | Scenario A | Scenario B | Scenario C | Reference Scenario | Scenario A | Scenario B | Scenario C |
| **Emergency Department Visits** |  |  |  |  |  |  |  |  |
| <6 months | 1.0 | 72.6 (57.6-77.6), | 65.8 (51.0-72.1) | 41.4 (32.1-45.3) | Referent | 262 (211-301) | 238 (187-277) | 150 (118-174) |
| 6–11 months | 1.0 | 34.7 (25.4-41.9) | 27.7 (20.2-33.6) | 17.6 (12.9-21.4) | Referent | 37 (27-46) | 30 (22-37) | 19 (14-24) |
| All ages | 1.0 | 23.9 (19.7-26.4) | 21.7 (17.7-24.2) | 11.9 (9.5-13.4) |  |  |  |  |
| **Hospitalizations** |  |  |  |  |  |  |  |  |
| <6 months | 1.0 | 73.4 (59.0-77.6) | 66.7 (52.3-72.3) | 41.9 (32.9-45.4) | Referent | 85 (70-97) | 77 (62-89) | 49 (39-56) |
| 6–11 months | 1.0 | 35.5 (26.1-42.4) | 28.3 (20.8-34.1) | 18.0 (13.2-21.7) | Referent | 12 (9-15) | 10 (7-12) | 6 (5-8) |
| All ages | 1.0 | 30.7 (25.8-32.9) | 28.3 (23.6-30.7) | 14.0 (11.2-15.3) |  |  |  |  |

Abbreviations: UI, uncertainty interval. \*Scenarios described in Table 1.

## Discussion

Through the applied use of an accessible modeling tool, this analysis using routine public health surveillance data estimated the state-level impact of new RSV immunization products for at-risk groups. When compared to a reference scenario in which no new immunizations were administered, the youngest at-risk age groups showed significantly lower ED visits and hospitalizations during the 2024–2025 season for both highly immunized infant populations as well as a scenario reflecting actual 2024–2025 immunization coverage. By modeling a “typical” RSV season against multiple possible immunization coverage scenarios, this analysis allowed for further exploration of the impact of different immunization products and coverage levels of vaccination for vulnerable populations to inform public health action and communication. In a typical RSV season, immunizing nearly every infant—either at birth, through a catch-up dose, or via conferred immunity through vaccination during pregnancy—could result in 72.6% lower ED visits and 73.4% lower hospitalizations for RSV, or 262 ED visits and 85 hospitalizations averted per 10,000 population. Projected ED visits and hospitalizations were also lower among the 6–11-month-old age group, although only a portion of the cohort is eligible for immunization. While these coverage scenarios were all achievable during the 2024–2025 season, during periods of supply constraint, tools like this could help inform recommendations for optimal immunization allocation and administration, including by further stratifying modeling outputs by at-risk demographics. The findings from this analysis help contextualize the strong efficacy of both monoclonal antibody administration and vaccination during pregnancy for preventing severe illness in infants in Massachusetts.

Public health surveillance data can be used to quantify the burden of RSV infection and severe illness but is often unable to be linked to other key data sources, like immunization data, to assess the impact of public health interventions. Other limitations inherent to RSV surveillance data, like disruptions to care seeking behavior, changes in RSV testing practices, shifts in disease patterns, and practical considerations like the availability of data, made RSV an ideal candidate for the use of an off-the-shelf modeling tool to generate insights into impacts on the burden and transmission of RSV. While scenario projections are often used prospectively to explore the impact of public health interventions in advance of respiratory seasons [25], these types of models can also be utilized during seasonal epidemics or public health emergencies to better understand and contextualize the impact of interventions. Mathematical and statistical models are built on key assumptions like infectious disease dynamics, population characteristics, and intervention effectiveness, and include measures of uncertainty to provide a realistic view of possible outcomes. As such, model outputs should be framed and communicated alongside these key assumptions.

While the total observed hospital encounters for the 2024–2025 season were higher than the median scenario projections, review of the 2024–2025 RSV season showed overall higher intensity as compared to past seasons, lending helpful context to the model estimates. Applying practical, methodological, and evidence-based adjustments to historical data inputs and assessing model outputs against other information from the public health toolbox—like seasonal intensity thresholds [26,27] and correlation with observed data—can yield helpful situational insights for public health decision-makers.

The advancement of modeling tools for applied public health at the STLT level has historically been accomplished through close collaboration with academic partners, often including the wholesale outsourcing of analyses to modelers which can cause separation between academic expertise and practical applied public health work. However, the increasing availability of accessible, off-the-shelf modeling tools supports ease of adoption by STLT jurisdictions, particularly when paired with consultation from academic modeling partners. Scaling these tools within a public health department can be especially effective with close collaboration across epidemiological program areas to build recognition and inform model inputs. These advanced tools allow for innovations in public health surveillance by estimating and communicating public health impact in new and different ways. Despite the increasing availability and accessibility of readily available infectious disease modeling tools, sustained investment and resources are still required to support the public health workforce upskilling and operational activities necessary to facilitate the use of these advanced analytic methods.

### Limitations

There are some limitations to the findings of this report. Syndromic surveillance hospital encounter data were categorized as RSV-related based on a syndrome definition that utilizes chief complaint and discharge diagnosis as recorded in the electronic health record rather than laboratory diagnosis. Additionally, the approach to scaling hospital encounter volume to account for changes in testing and administrative coding behavior in addition to missing facility volume may not reflect the true burden of RSV associated encounters. The modeling tool includes fixed susceptibility and infectiousness parameters informed by the literature and may not reflect precise real-world transmission dynamics or immunization efficacy. Additionally, the tool assumes constant immunization administration within the population throughout the season and may therefore underestimate the impact of vaccination among infant populations. This analysis included hospital encounter data for patients with Massachusetts addresses only, and utilized 2022 census data, which may not be representative of the full susceptible population for the 2024–2025 season.

## Conclusions

Analysis of routinely available public health surveillance data using this model was both practical and useful to inform applied public health practice. Estimation of the impact of RSV immunization on hospital encounters using this type of modeling tool provides important public health insights. The results suggest that RSV immunizations for infant populations can result in significantly lower RSV-related ED visits and hospitalizations in Massachusetts. Scenario projections for the 2024–2025 season indicate that immunizing most eligible infants under 6 months of age could result in 72.6% lower ED visits and 73.4% lower hospitalizations for RSV than if no infants were immunized. Public health workforce upskilling and resource investment are necessary for implementing readily accessible off-the-shelf infectious disease modeling tools.

## Acknowledgments

We thank Kathryn Ahnger-Pier, Pejman Talebian, Spencer Cunningham, Petra Schubert, Mia Haddad, Larry Madoff, Angela Fowler, and Meagan Burns for their valuable feedback. We thank Chelsea Hansen for development of the modeling tool.

## Funding Statement

The authors acknowledge financial support from CDC Grant NU38FT00008 (L.J., R.E., A.T., C.B., N.G.R., R.P.).

## Conflicts of Interest

N.G.R. discloses a consulting relationship with Google LLC, although this funding and research are unrelated to the current publication. All other authors declare no conflicts of interest.

## Data Availability

Example data for the model explored in this manuscript is available at: https://github.com/chelsea-hansen/R.Scenario.Vax/ [10]. An application for data access to the aggregate time series data utilized for this analysis can be made to the Massachusetts Department of Public Health: https://www.mass.gov/lists/infectious-disease-data-reports-and-requests.

## Ethics Approval

The Institutional Review Board (IRB) of the Massachusetts Department of Public Health (DPH) waived ethical approval for this work. The Massachusetts DPH IRB follows federal regulations regarding ethical review. All data for this analysis were collected through routine public health surveillance activities as per Federal Regulation 45 CFR 164.512(b). These regulations authorize public health authorities to collect data for disease control, including reporting disease, vital events, and conducting public health investigations. According to the Common Rule (Federal Regulation 45 CFR 46 subpart A §46.104), data collected for public health purposes are exempt from IRB review, as detailed in Paragraph D5. Furthermore, research conducted by federal agencies using government-collected information for non-research activities is also exempt, provided that identifiable private information is maintained in compliance with the E-Government Act of 2002, the Privacy Act of 1974, and, when applicable, the Paperwork Reduction Act of 1995. This ensures the data remain protected while serving public health objectives. Informed consent was not required because all data were collected through routine public health surveillance activities as per Federal Regulation 45 CFR 164.512(b).

## Author Contributions

Conceptualization: Laura Jones, Rosa Ergas, Andrew Tibbs

Data curation: Laura Jones, Josh Norville, Boudu Bingay

Formal analysis: Laura Jones

Funding acquisition: Catherine M. Brown, Nicholas G. Reich

Investigation: Laura Jones, Josh Norville, Boudu Bingay

Methodology: Laura Jones, Rosa Ergas, Andrew Tibbs, Elizabeth T. Russo, Catherine M. Brown

Project administration: Laura Jones

Resources: Catherine M. Brown Software: Laura Jones

Supervision: Catherine M. Brown, Nicholas G. Reich, Remy Pasco

Validation: Laura Jones, Rosa Ergas, Andrew Tibbs

Visualization: Laura Jones

Writing – original draft: Laura Jones

Writing – review & editing: Laura Jones, Rosa Ergas, Andrew Tibbs, Elizabeth T. Russo, Josh Norville, Boudu Bingay, Catherine M. Brown, Nicholas G. Reich, Remy Pasco

## Abbreviations

RSV: Respiratory Syncytial Virus
ED: Emergency Department
DPH: Massachusetts Department of Public Health
NSSP: National Syndromic Surveillance Program
MIIS: Massachusetts Immunization Information System
STLT: State, tribal, local, and territorial
PI: Projection Interval
UI: Uncertainty Interval

## Notes

### Author Declarations

The Institutional Review Board (IRB) of the Massachusetts Department of Public Health (DPH) waived ethical approval for this work. The Massachusetts DPH IRB follows federal regulations regarding ethical review. All data for this analysis were collected through routine public health surveillance activities as per Federal Regulation 45 CFR 164.512(b). These regulations authorize public health authorities to collect data for disease control, including reporting disease, vital events, and conducting public health investigations. According to the Common Rule (Federal Regulation 45 CFR 46 subpart A, Section 46.104), data collected for public health purposes are exempt from IRB review, as detailed in Paragraph D5. Furthermore, research conducted by federal agencies using government-collected information for non-research activities is also exempt, provided that identifiable private information is maintained in compliance with the E-Government Act of 2002, the Privacy Act of 1974, and, when applicable, the Paperwork Reduction Act of 1995. This ensures the data remain protected while serving public health objectives. Informed consent was not required because all data were collected through routine public health surveillance activities as per Federal Regulation 45 CFR 164.512(b).

