## Supplement for "Modeling the Impact of Pediatric RSV Immunization in Massachusetts, 2024–2025"

### *DPH RSV Immunization Recommendations at Time of Analysis*

At the time of analysis, the RSV vaccine was recommended for pregnant people at 32 through 36 weeks of pregnancy during the months of September through January. Monoclonal antibody administration of *nirsevimab* was recommended for infants less than 8 months old from October 1 through March 31 whose birth parent did not receive the RSV vaccine during pregnancy.

### *Historical Emergency Department Encounter Data Scaling Approach*

**Supplement Table 1.** Historical RSV Emergency Department encounter data facility scaling, Massachusetts, United States. Historical data prior to January 1, 2019, were initially scaled to account for the missing encounter volume from a large hospital system not yet enrolled in the syndromic surveillance program.

| | Proportion of 2019–2020 statewide RSV visits | |
|---|---|---|
| Age group | Emergency Department Visits | Hospitalizations |
| <6 months | 0.2099 | 0.2207 |
| 6–11 months | 0.2176 | 0.2851 |
| 1–4 years | 0.2405 | 0.2368 |
| 5–64 years | 0.3162 | 0.3210 |
| 65–74 years | 0.2562 | 0.2836 |
| 75+ years | 0.2901 | 0.2679 |

**Supplement Table 2**. Historical RSV Emergency Department encounter data post-pandemic scaling, Massachusetts, United States. Historical data were further scaled by the change in volume between 2018–2019 and 2023–2024 RSV seasons by age group [1,2].

| | Post-pandemic scaling factor | |
|---|---|---|
| Age group | Emergency Department Visits | Hospitalizations |

| | | |
|---|---|---|
| <6 months | 1.2x | 1.2x |
| 6–11 months | 1.6x | 1.3x |
| 1–4 years | 2.3x | 2.1x |
| 5–64 years | 2.3x | 2.1x |
| 65–74 years | 2.3x | 2.1x |
| 75+ years | 2.3x | 2.1x |

**Supplement Table 3**. Historical RSV Emergency Department encounter data pre-pandemic scaling, Massachusetts, United States. The 2017–2018 and 2018–2019 season historical data were also scaled to account for year-over-year increase in respiratory testing practices observed prior to the COVID-19 pandemic [3].

| | Pre-pandemic scaling, slope | |
|---|---|---|
| Age group | Emergency Department Visits | Hospitalizations |
| All ages | 0.25 | 0.15 |

***Historical Emergency Department Encounter Calculated Age Distribution***

To represent a "typical" season's encounter burden by age group, overall age distribution for both ED visits and hospitalizations, were calculated using the average proportion of encounters by age group from the prior two RSV seasons (2022–2023 and 2023–2024) to account for the most recent trends in age of patients with ED visits or hospitalizations for RSV.

**Supplement Table 4**. Average Age Distribution of Emergency Department Encounters, 2022–2023 and 2023–2024 RSV Seasons, Massachusetts, United States.

| Age Group | Emergency Department Visits | Hospitalizations |
|---|---|---|
| <6 months | 0.1862604 | 0.24752901 |
| 6–11 months | 0.13107806 | 0.09626128 |
| 1–4 years | 0.35384677 | 0.21443919 |
| 5–64 years | 0.20037588 | 0.1486893 |
| 65–74 years | 0.04678503 | 0.08881249 |

| | | |
|---|---|---|
| 75+ years | 0.08165387 | 0.20426873 |

### ***Fixed Model Parameters***

Fixed susceptibility and infectiousness parameters based on published literature [4-12] were defined by the R.Scenario.Vax modeling tool developer [13] and were utilized for this analysis.

**Supplement Table 5.** Fixed RSV Scenario Model Parameters

| Parameter | Value |
|---|---|
| Duration of infectiousness - first infection ($1/\gamma_1$) [28] | 10 days |
| Duration of infectiousness - second infection ($1/\gamma_2$) | 7 days |
| Duration of infectiousness - third or later infection ($1/\gamma_3$) | 5 days |
| Relative risk of infection following first infection ($\sigma_1$) | 0.89 |
| Relative risk of infection following second infection ($\sigma_2$) | 0.72 |
| Relative risk of infection following third or later infection ($\sigma_3$) | 0.24 |
| Relative risk of infection with maternal immunity (same as RR following third infection) ($\sigma_3$) | 0.24 |
| Duration of maternal immunity ($1/\omega_1$) | 90 days |
| Duration of immunity following first and second infections ($1/\omega_2$) | 182.625 days |
| Duration of immunity following third or later infections ($1/\omega_3$) | 358.9 days |
| Relative infectiousness - second infections ($\rho_1$) | 0.75 |
| Relative infectiousness - third or later infections ($\rho_2$) | 0.51 |
| Baseline transmission rate ($\beta$) | Fitted |
| Amplitude of seasonal forcing ($b_1$) | Fitted |
| Phase of seasonal forcing ($\varphi$) | Fitted |
| Infections that lead to reported hospitalizations (<2m, 2-11 months fixed relative to this) | Fitted |
| Infections that lead to reported hospitalizations (1-4 yrs) | Fitted |
| Infections that lead to reported hospitalizations (5-64 yrs) | Fitted |

| Parameter | Value |
|---|---|
| Infections that lead to reported hospitalizations (65-74 yrs) | Fitted |
| Infections that lead to reported hospitalizations (75+ yrs) | Fitted |
| Birth rate for newborns who are not protected by immunizations (B1) | Scenario Based |
| Birth rate for newborns protected by monoclonal antibodies (B2) | Scenario Based |
| Birth rate for newborns protected by maternal vaccination (B3) | Scenario Based |
| Catch-up immunization rate for monoclonal antibodies (U) | Scenario Based |
| Vaccination rate for older adults (V) | Scenario Based |
| *Nirsevimab* effectiveness | 80% |
| Maternal vaccination effectiveness | 57% |
| Duration of infant immunizations (both monoclonals and maternal vaccination) $2*(1/\omega_i)$ | 180 days |
| Vaccine effectiveness in older adults | 75% |
| Duration of vaccine effectiveness in older adults $2*(1/\omega_v)$ | 2 years |

***Comparison to 2024-2025 Observed Hospital Encounters***

**Supplement Figure 1.** Median Scenario Projections vs. Observed RSV Hospital Encounters, 2024–2025 Season, Massachusetts, United States. Scenarios described in Manuscript Table 1.

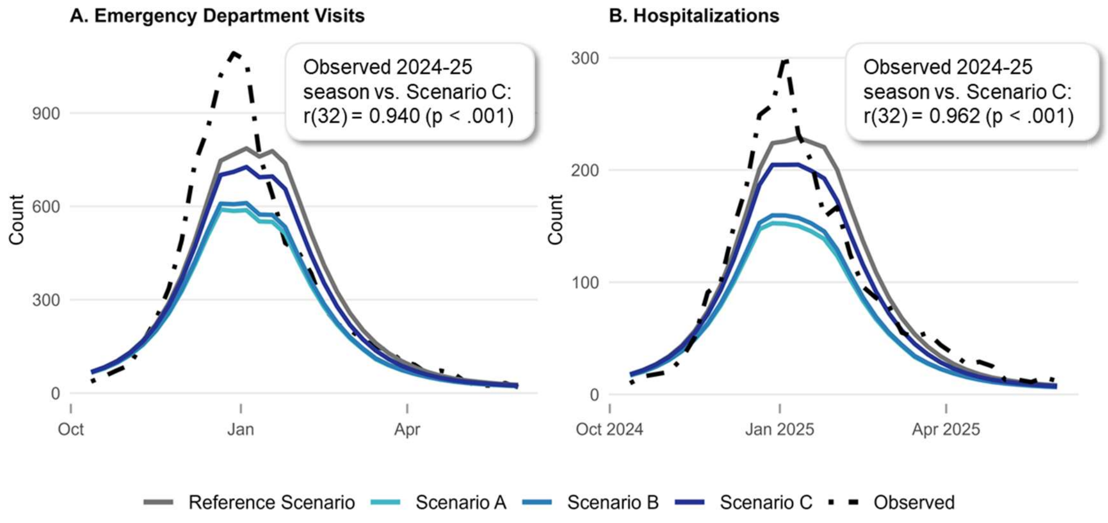
